# Pilot Validation of an AI-based Audiovisual Fatigue Assessment Tool (mAI Fatigue) in Chronic Liver Disease: A Multicentre Study

**DOI:** 10.64898/2026.06.22.26356228

**Authors:** Gourdas Choudhuri, Gamar Akhundova-Unadkat, Ana Maria Rodriguez-Leboeu, Michel Valstar, Kinjalkumar Shah, Ruben Duijnhoven, Azadeh Safaei, Mark G. Swain

**Affiliations:** Department of Gastroenterology and Hepato-Biliary Sciences, Fortis Memorial Research Institute, Gurugram, Haryana, India; Uerzlikon, Switzerland; IQVIA, Madrid, Spain; Blueskeye® Ltd, University of Nottingham, Nottingham, UK; Abbott Healthcare Pvt Ltd, Mumbai, India; Abbott Laboratories, Global Biometrics and Data Center, C.J. van Houtenlaan 36, 1381 CP Weesp, the Netherlands; Abbott Product Operations AG, Allschwil, Switzerland; University of Calgary, Calgary, Canada

**Author notes:** Corresponding author: Mark G. Swain; Calgary Liver Unit, Department of Medicine, Snyder Institute for Chronic Diseases, Cumming School of Medicine, University of Calgary, Calgary, AB, Canada.

## Abstract

Fatigue affects over half of patients with chronic liver disease (CLD) and is a major driver of impaired quality of life, yet it remains under-recognised because assessment relies almost entirely on subjective patient-reported outcomes (PROs). This proof-of-concept study evaluated whether audiovisual (AV) markers from facial and vocal expressions, captured via the mAI Fatigue tool (Blueskeye®), could serve as objective correlates of fatigue in CLD. In a prospective, multicentre, case-control study at three sites in India, 111 adults (aged 18–65 years) were enrolled as healthy controls (n=55) or CLD patients with moderate-to-severe fatigue (n=56). Over four weeks, participants completed ten assessments combining validated PROs, Psychomotor Vigilance Task (PVT) reaction times and AV recordings. CLD participants had significantly slower PVT reaction times than controls (882 vs 776 ms; p=0.0047). Session-level AV–PRO correlations were modest (r=−0.17 to −0.27), but participant-level aggregation strengthened associations (r=−0.47; p≈0.002) in the high-quality audio subset (n=41), where a predictive model achieved R=0.75–0.76 (p<0.001); associations were strongest in older participants, women, those with severe fatigue and MASLD aetiology. AI-derived AV markers, particularly when anchored to an individual baseline and aggregated longitudinally, show promise as objective, complementary measures of fatigue in CLD and warrant validation in larger, diverse cohorts.

**Clinical trials registration:** www.ctri.nic.in, CTRI/2024/12/077720.

## Introduction

Fatigue is a prevalent and debilitating symptom, affecting approximately 5–7% of the general population and over 50% of individuals with chronic liver disease (CLD)^1–3^. It significantly impairs health-related quality of life (HRQoL), with women experiencing more severe symptoms than men across all age groups^4^. Fatigue is characterised by reduced energy, persistent exhaustion unrelieved by rest, and a perceived inability to perform mental and physical activities^5^. It manifests as both central and peripheral symptoms, including cognitive decline, sleep disturbances, apathy, autonomic dysfunction, muscle fatigue, and decreased exercise capacity^1–3^.

Central fatigue, often exacerbated by liver inflammation in CLD patients, is associated with changes in brain neurotransmission, while peripheral fatigue stems from neuromuscular dysfunction^2,6^. In metabolic dysfunction-associated steatotic liver disease (MASLD), fatigue is the most common complaint and a key driver of reduced HRQoL^7^. Reported prevalence among patients with CLD varies widely, from 45% to 85% depending on methodology, aetiology, and disease stage^8^. In MASLD specifically, fatigue is among the most clinically significant symptoms, with severity classified as severe in 36–49% of patients and shown to exceed levels in the general population in an international registry of 5,691 patients^8–11^. It is linked to social dysfunction, daytime sleepiness, reduced work capability, and increased mortality risk^1,12–15^.

Despite its high prevalence and impact, fatigue remains under-recognised, under-diagnosed, and inadequately managed due to the lack of reliable and objective assessment tools^3^. Integrating machine learning with established patient-reported outcome (PRO) instruments would address the limitations of purely subjective methods and support improved fatigue identification^16,17^.

Recent advancements in machine learning offer promising opportunities to develop objective methods for assessing fatigue in chronic diseases. By analysing complex datasets, machine learning models can identify patterns that traditional methods may overlook, providing a more comprehensive understanding of fatigue intensity^18^. The mAI Fatigue tool, developed by Blueskeye® (https://www.blueskeye.com/b-healthy/; accessed 17 April 2026) in 2023, represents a novel approach to fatigue assessment. This software development kit uses machine learning algorithms to analyse facial and vocal data, offering objective insights into expressed emotions, discomfort, stress, and fatigue. By measuring behavioural cues such as facial muscle activation, gaze, and voice-acoustic features, the tool aims to provide a more accurate and reliable assessment of fatigue compared with subjective measures, e.g., existing PROs.

This proof-of-concept study aimed to evaluate audiovisual (AV) parameters derived from facial and vocal expressions as potential objective markers of fatigue intensity in adults with CLD and moderate-to-severe fatigue, compared with a healthy population with no fatigue (non-fatigued controls). Secondary aims were to examine the correlation between AV markers and validated PRO measures, and to develop a predictive regression model for fatigue assessment. By integrating machine learning with established PRO instruments, we sought to address the limitations of purely subjective methods and support improved fatigue identification and management in CLD.

## Results

### Participant disposition and baseline characteristics

Of the 116 participants who provided informed consent, 111 were enrolled in the study, and 100 completed with analysable longitudinal data. Five participants were excluded due to screen failures. Cohort A (non-fatigued controls) included 55 participants, of whom 54 (98.2%) completed the study. Cohort B (CLD with moderate-to-severe fatigue) included 56 participants, with 50 (89.3%) completing the study. Reasons for premature termination in Cohort B included loss to follow-up (n=3), withdrawal of consent (n=2), and protocol violation (n=1). A detailed flowchart of participant disposition and completion rates are presented in **Supplementary Material_Figure S2**, and **Table S2**, respectively.

The Asian study population was predominantly male (79.3%), with female participants comprising the remaining 20.7%. The mean age was 36.6 years (SD 9.34), with most participants (39.6%) aged 30–39 years. Cohort B participants were diagnosed with CLD, with the majority (58.9%) having MASLD; other CLD aetiologies included alcoholic liver disease (28.6%), viral hepatitis E (7.1%) and C (1.8%), non-alcoholic steatohepatitis (1.8%) and primary biliary cholangitis (1.8%) (**Supplementary Material_Table S3**).

None of the participants in Cohort A reported any medical history. In Cohort B, 14 participants (25.0%) reported at least one medical condition, with diabetes mellitus being the most common (21.2%). Other reported conditions included hypertension (7.1%) and dyslipidaemia (1.8%). Baseline demographic and clinical characteristics are summarised in **Table 1**.

**Table 1.** Baseline patient demographics and characteristics.

| Characteristics | Cohort A<br>(N=55) | Cohort B<br>(N=56) | All Participants<br>(N=111) |
| --- | --- | --- | --- |
| Age (years), mean (SD) | 36.8 (9.44) | 36.4 (9.33) | 36.6 (9.34) |
| **Age Category (yrs), n (%)** |  |  |  |
| 18 to 29 | 17 (30.9) | 14 (25.0) | 31 (27.9) |
| 30 to 39 | 20 (36.4) | 24 (42.9) | 44 (39.6) |
| 40 to 49 | 13 (23.6) | 10 (17.9) | 23 (20.7) |
| 50 to 59, | 4 (7.3) | 8 (14.3) | 12 (10.8) |
| ≥60 | 1 (1.8) | 0 | 1 (0.9) |
| **Gender, n (%)** |  |  |  |
| Male | 43 (78.2) | 45 (80.4) | 88 (79.3) |
| Female | 12 (21.8) | 11 (19.6) | 23 (20.7) |
| **Race, n (%)** |  |  |  |
| Asian | 55 (100) | 56 (100) | 111 (100) |

standard
deviation.

### Reaction time analysis

The Psychomotor Vigilance Task (PVT) is a brief, validated reaction-time test in which participants press a key in response to a visual stimulus appearing at random intervals; median PVT reaction time is widely used as an objective behavioural index of fatigue-related cognitive slowing and was used here as the per-session objective fatigue anchor against which AV markers were evaluated. The study demonstrated that the CLD group had a statistically significant slower median reaction time on the PVT compared to the Control group, with the CLD group showing a median reaction time of 882 ms versus 776 ms in the Control group (p=0.0047) (**Fig. 1**).

**Figure 1.**
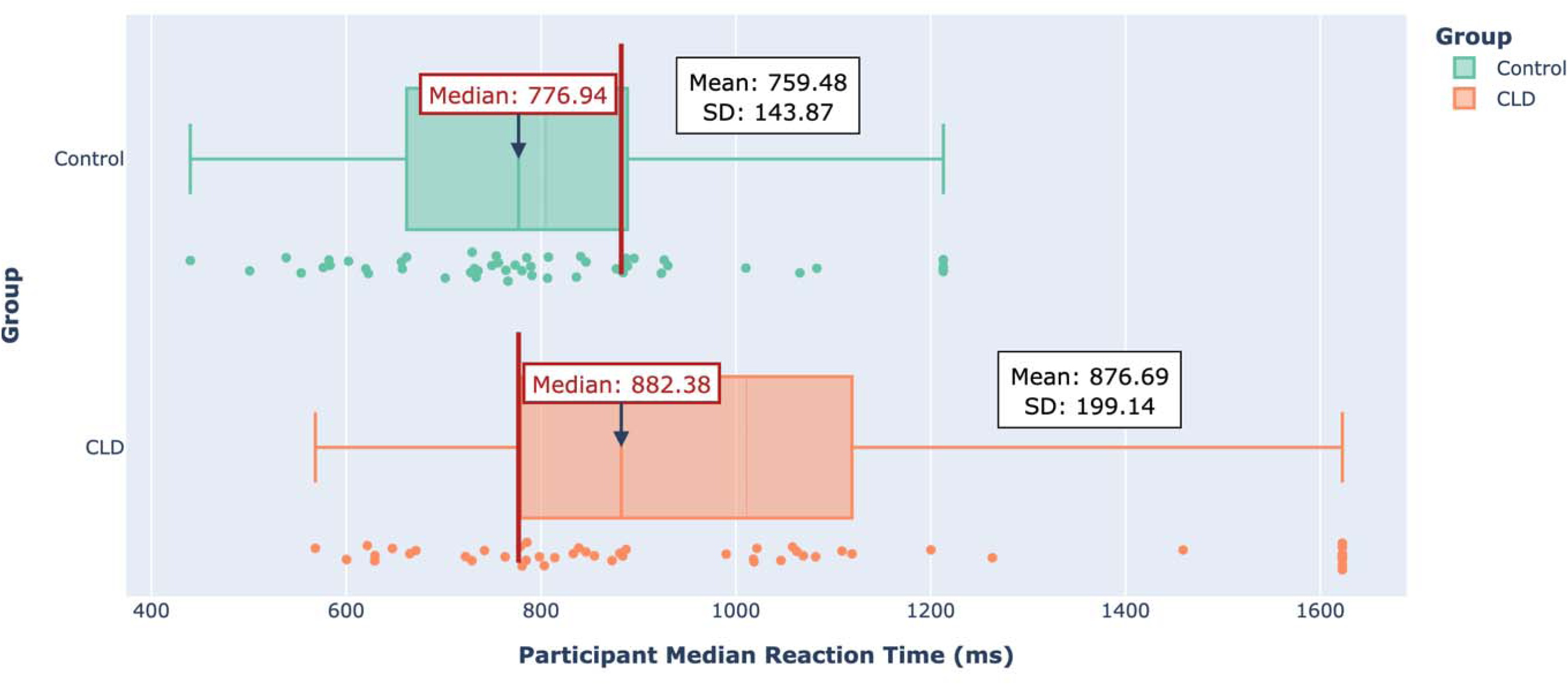
Comparison of per participant median reaction time (across all visits) between CLD and Control groups CLD, chronic liver disease; ms, milliseconds; SD, standard deviation.

### Longitudinal tracking of AV fatigue markers

Longitudinal tracking refers here to the within-participant assessment of AV fatigue markers across the 10 study sessions over four weeks, which allowed each participant to serve as their own baseline and enabled detection of trend-level changes that single-session measurements cannot capture. AV fatigue markers are computer-vision and voice-analysis features extracted by the Blueskeye® platform from short smartphone-recorded video and read-aloud / picture-description audio clips, and span five categories — blink dynamics, gaze behaviour, voice acoustics, facial action units (AUs), and mouth movements; the full marker list is provided in **Supplementary Material_Table S4**.

After filtering high-quality data, a significant number of robust markers emerged; ‘high-quality’ refers to AV recordings where factors, e.g., face visibility, eye visibility, head pose range, sound quality, speech fluency and task adherence, were rated as good during a manual annotation process. Filtering by quality was necessary due to a high volume of low-quality AV recordings and subsequent potentially unreliable interpretation of fatigue biomarkers; manual audiovisual quality annotation results for each task are summarised in **Supplementary Material_Table S5**. Markers were deemed reliable if at least 60.0% of participants exhibited a consistent directional trend (positive or negative slope). In the audio quality subset, 26 markers met this threshold (e.g., Blink: 4, Gaze: 5, Voice: 6, AUs: 9, Mouth: 2), while in the visual quality subset, 28 markers met the threshold (e.g., Blink: 11, Gaze: 5, Voice: 5, AUs: 7, Mouth: 0); individual markers are listed in **Supplementary Material_Table S6**.

Inter-participant variability was clear in both baseline values and response magnitudes. Baseline variation showed wide differences in starting points for low-fatigue states, while magnitude variation highlighted significant differences in how much marker values changed between participants. **Supplementary Material_Figure S3** illustrates these trends, showing examples of strong positive trends (e.g., Audio Metric 4, Eye Lid Metric 1), strong negative trends (e.g., Audio Metric 20), and variable trends (e.g., Gaze Metric 2).

The analysis also demonstrated strong intra-participant correlations, confirming that AV biomarkers effectively tracked fatigue over time. Individual tracking performance analysis revealed that approximately 72.0% of participants with at least three sessions showed a strong longitudinal correlation between their PVT scores and at least one AV fatigue marker (**Fig. 2**). On a cohort-wide level, 72.0% of participants showed consistent patterns across the 10 sessions over four weeks (Trend Coherence Score > 0.6) between their PVT performance and at least one AV fatigue marker. Filtering for data quality improved these results, with 84.9% of participants with high-quality audio data and 78.1% with high-quality visual data showing strong alignment.

**Figure 2.**
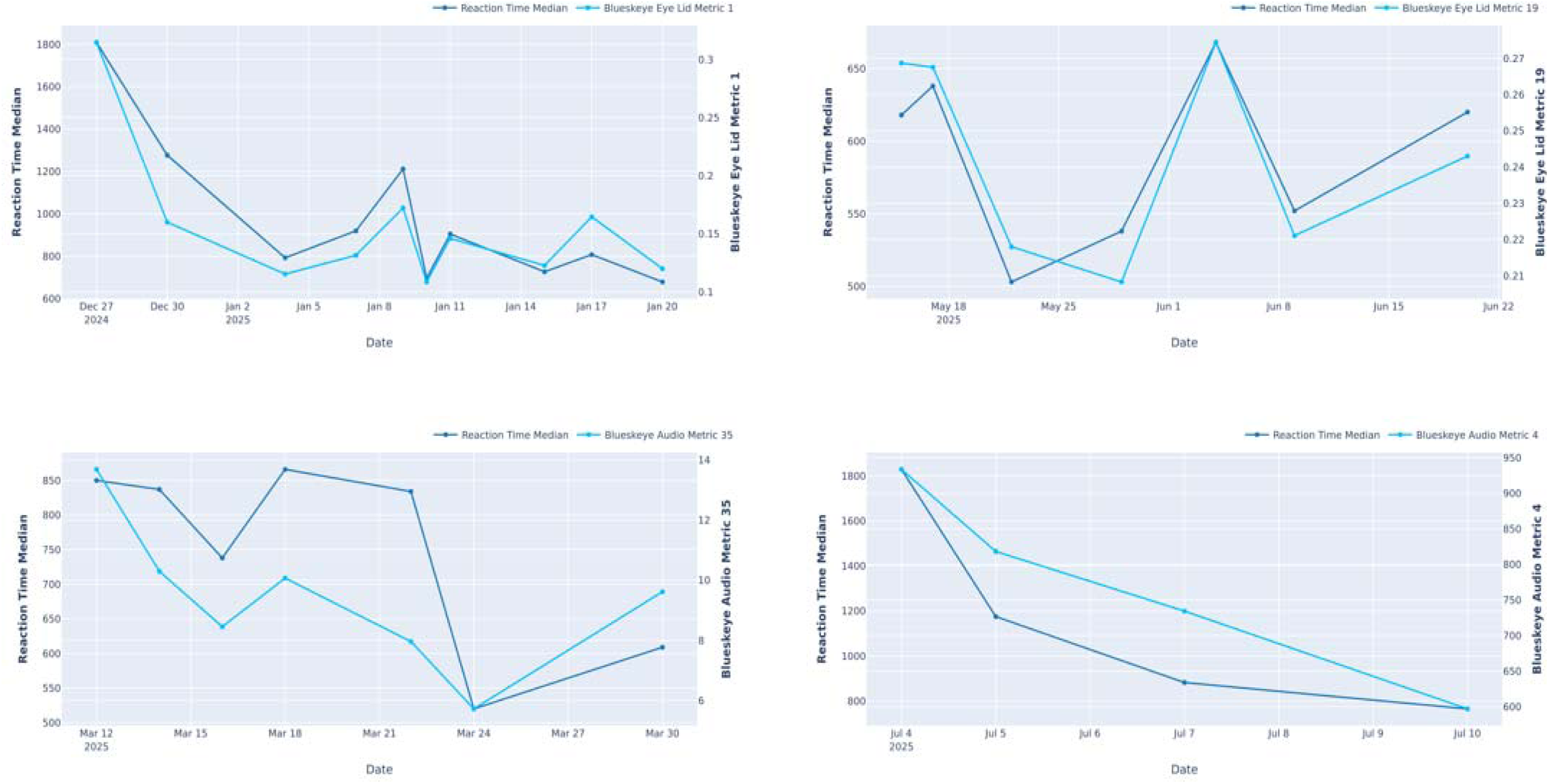
Examples of strong intra-participant (within-subject) longitudinal correlations Each plot tracks a participant’s objective fatigue (reaction_time_median, dark blue) and a specific AV biomarker (light blue) over multiple sessions, demonstrating the marker’s high sensitivity to individual fatigue changes over time.

### Correlation analysis between AV metrics and PROs

Session-level correlations between the top 5 ranked AV fatigue markers and PRO scores (MFI Physical Fatigue and MFI Mental Fatigue) were statistically significant but modest, with correlation coefficients ranging from r=–0.17 to –0.27 (p<0.05). However, participant-level temporal aggregation of AV data markedly strengthened the associations between AV metrics and participants’ chronic fatigue (PRO) scores, which were absent in the session-level analysis. In the high-quality audio subset (n=41), strong negative correlations were observed between AV metrics and PRO fatigue scores, with correlations of r = –0.47 (p=0.0019) for physical fatigue and r = –0.47 (p=0.0021) for mental fatigue. In the high-quality visual subset (n=55), moderate correlations were found, with r = 0.35 (p=0.0097) for physical fatigue and r = 0.35 (p=0.0097) for mental fatigue (**Supplementary Material_Figures S4 and S5**, respectively).

### Correlation of AV markers and fatigue scores by subgroup

#### Age

Correlations between AV markers and fatigue scores varied by age group. Younger participants (18–29 years) showed moderate negative correlations between AV markers and physical fatigue, particularly in Eye Tracking (–0.46; p=0.053)) and Picture Description (–0.40; p=0.0002) tasks. Younger participants (18–29 years) did not show any notable positive correlations. Participants aged 30–39 years also showed moderate correlations, with markers linked to mental fatigue in Eye Tracking (–0.32; p=0.031) and physical fatigue in Read-Aloud (–0.30; p=0.0003).

For participants aged 40–49 years, stronger correlations were observed, with a positive correlation of 0.46 in Eye Tracking (p<0.0001) and a negative correlation of –0.44 in Picture Description (p=0.0008), both linked to physical fatigue. The strongest correlations overall were seen in participants aged 50–59 years, with markers showing –0.76 for mental fatigue in Eye Tracking (p=0.006) and –0.69 for physical fatigue in Picture Description (p=0.0001). No meaningful analysis was possible for participants aged 60 and older due to a small sample size.

#### Gender

Significant differences in how AV markers correlated with fatigue scores between males and females were observed. For male participants, moderate correlations were observed in the Read-Aloud task, with markers showing –0.28 (p<0.0001) and –0.25 for physical fatigue (p=0.0003). For female participants, stronger correlations were observed than in males (peak r = 0.43 vs −0.28). In the Read-Aloud task, a marker showed a positive correlation of 0.43 with mental fatigue (p<0.0001), while in the Picture Description task, another marker showed –0.40 with mental fatigue (p=0.0008).

#### Fatigue severity

Analysis by fatigue severity showed that AV markers were more effective in tracking fatigue as its severity increased. For participants with moderate fatigue, a marker in the Picture Description task showed a correlation of –0.47 with physical fatigue (p=0.0003). In participants with severe fatigue, the same marker demonstrated a stronger correlation of –0.55 with physical fatigue (p=0.021; **Supplementary Material_Table S7**).

#### CLD aetiology

The analysis of participants with different types of CLD revealed varying correlations between AV markers and fatigue scores. For participants with viral hepatitis E, one marker appeared to show perfect correlations (r = 1.0) with both physical and mental fatigue; however, with only four participants in this subgroup, this result is almost certainly an artefact of the small sample and should be regarded as hypothesis-generating rather than a genuine effect.

In participants with MASLD, moderate correlations were observed. A marker in the Picture Description task showed –0.41 with physical fatigue (p=0.018), while another in the Eye Tracking task showed –0.41 with mental fatigue (p=0.056). For participants with other CLD aetiologies, the strongest correlations were observed in the Eye Tracking task, with markers showing 0.55 for physical fatigue (p=0.019) and 0.54 for mental fatigue (p=0.020).

### Evaluation of AV markers for fatigue prediction

In this context, ‘fatigue prediction’ refers to using AV markers as inputs to regression models that estimate a participant’s PRO-derived fatigue score (i.e., model-predicted vs PRO fatigue intensity), rather than temporal forecasting of future fatigue states. The secondary analysis demonstrated that AV markers can predict fatigue levels, with performance improving significantly when high-quality data subsets were used. In the full dataset, the models showed weak but statistically significant correlations for physical fatigue (R = 0.2814, p<0.05) and mental fatigue (R = 0.3024, p<0.05). However, in the good audio quality subset (N=41), the models achieved strong correlations for both physical fatigue (R = 0.7501, p<0.001; **Supplementary Material_Figure S6**) and mental fatigue (R = 0.7613, p<0.001). Similarly, the good visual quality subset showed moderate correlations, with mental fatigue (R = 0.5344, p<0.001) outperforming physical fatigue (R = 0.3850, p<0.01; **Supplementary Material_Table S8 and Figures S7, S8, S9 and S10**).

Aggregating participant-level data over time further enhanced predictive accuracy, with models achieving stronger correlations compared to session-level analyses. For example, in the good audio quality subset, mental fatigue predictions reached a Pearson correlation of R = 0.7613 (p<0.001) with an RMSE of 0.2906, while physical fatigue predictions achieved R = 0.7501 (p<0.001) with an RMSE of 0.3005.

## Discussion

This proof-of-concept study highlights the potential of AI-derived AV markers, based on facial and vocal expressions, as sensitive and objective measures of fatigue in adults with CLD. By leveraging machine learning and longitudinal data aggregation, the findings of this study demonstrate that AV markers can effectively track fatigue fluctuations and complement traditional PROs.

The study successfully showcased the utility of Blueskeye® facial and vocal fatigue markers for objectively assessing fatigue through two distinct mechanisms. Temporal variability in AV markers, such as changes in voice and blink metrics across multiple sessions, showed moderate-to-strong correlations with chronic fatigue scores, suggesting that baseline-anchored digital phenotyping offers a robust and personalised approach to fatigue assessment. Additionally, AV markers were highly sensitive to real-time fatigue changes, as evidenced by strong correlations with PVT reaction times.

The results underscore the importance of high-quality longitudinal data and temporal features in maximising the sensitivity and reliability of AV markers. These findings align with previous research demonstrating the utility of machine learning in analysing complex datasets to identify patterns that traditional methods may overlook^19,20^. Notably, the proportion of participants meeting the high-quality threshold was higher for audio (84.9%) than for visual (78.1%) data, likely reflecting the greater robustness of audio capture to variation in lighting, framing, and participant positioning during smartphone-based assessments.

Subgroup analyses revealed variations in the relationship between AV markers and fatigue scores, with older participants, particularly those aged 50–59 years, showing the strongest correlations. This is consistent with evidence suggesting that fatigue expression and perception may vary with age due to differences in physiological and psychological resilience^21,22^. Gender differences were also observed, with female participants demonstrating stronger associations between AV markers and fatigue scores compared to males, reflecting prior findings that women often report more severe fatigue symptoms and may exhibit distinct behavioural and emotional responses to fatigue^4,21,23^.

Furthermore, the effectiveness of AV markers increased with fatigue severity, and variations were noted across different CLD aetiologies, with the strongest associations observed in participants with MASLD; apparent associations in the viral hepatitis E subgroup were based on only four participants and should be interpreted with caution. These findings suggest that AV-based fatigue assessment could be tailored to individual characteristics, such as age, gender, and disease type, to enhance its clinical utility.

Several strengths of this study are worth noting. To our knowledge, this is the first multicentre, prospectively designed evaluation of an AI-based audiovisual fatigue assessment tool in CLD, combining facial expression, vocal, gaze, and psychomotor (PVT) data streams within a single protocol. The longitudinal design, with ten assessments over four weeks, enabled both within-participant temporal aggregation and the use of personalised baselines — analytic approaches that are not feasible with one-off PRO administration and which proved critical to detecting robust correlations with fatigue. The use of four complementary, validated PRO instruments (MFI, FACIT-F, PROMIS-F, PGI-S) provided a multidimensional reference standard, and the inclusion of an objective behavioural anchor (PVT reaction time) allowed triangulation of AV markers against a non-self-report measure of fatigue-related cognitive slowing. Finally, enrolment across three Indian sites supports feasibility of remote, smartphone-based AV capture in a real-world, resource-variable setting

Despite its promising findings, the study had several limitations. The strongest results were derived from high-quality AV data subsets (n=41 for audio, n=55 for visual), with substantially weaker performance in the full dataset, highlighting the need for stringent quality control during data collection. Poor-quality data and inter-participant variability posed challenges for session-level analyses. Subgroup analyses by age, sex, severity, and aetiology were exploratory, were not pre-specified to be adequately powered, and were not adjusted for multiplicity; findings in the smallest strata (e.g., hepatitis E, n=4) should therefore be interpreted as hypothesis-generating. Fatigue status was defined by self-report (PGI-S) rather than by an objective clinical criterion, which may have introduced classification overlap between cohorts, and the four-week follow-up, while sufficient to demonstrate temporal aggregation effects, does not establish whether AV markers are sensitive to clinically meaningful change over longer horizons or in response to therapeutic intervention. In addition, and the relatively small sample size, particularly in subgroup analyses, limits the generalisability of the findings. The study population was also predominantly male (79.3%), relatively young (mean age 36.6 years), and recruited exclusively from three sites in India, further limiting generalisability to women, older adults, and other ethnic and healthcare settings. The CLD cohort was heterogeneous in aetiology, with MASLD predominating and some subgroups (e.g., viral hepatitis E, n=4) too small to support reliable inference; the perfect correlations observed in the smallest subgroups should be interpreted with caution. Larger validation studies are warranted to confirm these results and ensure their applicability across diverse populations.

Additionally, weak session-level correlations between AV markers and PRO scores may reflect partial construct overlap, as PROs primarily assess physical and functional aspects of fatigue, whereas AV markers may capture complementary dimensions, such as central or expressive fatigue.

The study also lacked measures targeting apathy, psychomotor slowing, or emotional expressivity, which could provide further insights into the behavioural dimensions of fatigue. Previous research has highlighted the importance of incorporating multidimensional measures to fully capture the complexity of fatigue^17,24^.

These findings support the use of AV-based, baseline-anchored digital phenotyping as a complementary method to PROs in clinical trials and potentially in clinical practice. By integrating AV markers into clinical workflows, this approach could enable personalised monitoring and early intervention in fatigue management. Future research should focus on validating these findings in larger cohorts, incorporating additional measures to capture a broader spectrum of fatigue-related dimensions, and developing real-time quality control mechanisms to improve data reliability.

Furthermore, exploring the integration of AV markers with other digital health technologies, such as wearable devices, could provide a more comprehensive and continuous assessment of fatigue^25,26^.

## Conclusions

Blueskeye®’s facial and vocal fatigue biomarkers show significant promise as tools for the longitudinal monitoring of behavioural and psychomotor correlates of fatigue, rather than as single-point, “one-off” assessments. The findings of this study emphasise the importance of establishing personalised baselines to effectively track in-the-moment fatigue fluctuations and leveraging temporal variability across multiple days to assess chronic fatigue states. By focusing on these priorities, future applications have the potential to provide more accurate, sensitive, and personalised insights into fatigue, paving the way for improved management and intervention strategies in both clinical and real-world settings.

## Methods

### Study design

This multicentre, prospective cohort study was conducted between December 2024 and August 2025 at three hospital-based sites in India: Aman Hospital and Research Centre (Vadodara, Gujarat), Renova Neelima Hospitals (Hyderabad, Telangana), and Sangvi Multispecialty Hospital Pvt. Ltd. (Pune, Maharashtra). The study aimed to evaluate audiovisual (AV) parameters derived from facial and vocal expressions as potential markers of fatigue (i.e., PRO scores collected for multiple types of fatigue: general, mental and physical, well-being, functional well-being, social/family well-being) in adults with CLD and moderate-to-severe fatigue, compared to a cohort of non-fatigued participants without CLD. The study was conducted in accordance with Good Clinical Practice (GCP) guidelines, the Declaration of Helsinki, and was approved by the Ethics Committees for all participating institutions: Aman Hospital and Research Center (ECR/857/Inst/GJ/2016/RR-19) on November 11, 2024; Neelima Hospitals (ECR/807/Inst/TG/2016/RR-24) on December 16, 2024; Sangvi Multispecialty Hospital (ECR/1865/Inst/MH/2023) on February 10, 2025. All participants provided written informed consent prior to enrolment.

### Study participants

Adults aged 18 to 65 years were eligible to participate if they provided written informed consent, including consent for AV recordings, and agreed to adhere to all study procedures. Participants were assigned to one of two cohorts. Cohort A included individuals without CLD who reported no fatigue, as assessed by the Patient Global Impression of Severity (PGI-S) tool with a response of “none.” Cohort B included individuals diagnosed with CLD who self-reported moderate-to-severe fatigue, as assessed by the PGI-S with responses of “moderate,” “severe,” or “very severe.”

Exclusion criteria included the lack of access to a compatible mobile device, recent treatment for fatigue (within three weeks prior to screening), Parkinson’s disease, or a history of alcohol or drug abuse within the three months prior to screening. Additionally, individuals with comorbidities or medical conditions (other than CLD or its related comorbidities) that could interfere with study participation were not eligible.

### Study Variables

The study’s primary endpoints were AV parameters linked to fatigue-related behavioural or psychomotor states. These included indicators such as reduced facial muscle tension, diminished expression intensity, drooping eyelids and mouth corners, tense lips, increased nasality, lower F1 and F2 frequencies, reduced fundamental frequency, slower speech articulation, and shifts in spectral energy distribution.

The secondary endpoints of the study involved developing a predictive regression model using Blueskeye® to map AV fatigue markers to normalised fatigue scores. Feature selection involved ranking AV markers by correlation with target scores and filtering for multicollinearity using variance inflation factor (VIF). Model performance was evaluated via root mean squared error (RMSE) and linear correlation analysis across combined cohorts.

Subgroup analyses examined correlations across predefined groups based on age, sex, fatigue severity (PGI-S), and CLD aetiology.

### Study procedures

Participants attended 10 study visits over a four-week period, including two site visits (screening/baseline and end-of-study [EOS]) and eight at-home assessments. At each visit, participants completed four validated fatigue assessment tools: the Multidimensional Fatigue Inventory (MFI), the Functional Assessment of Chronic Illness Therapy Fatigue Scale (FACIT-F), the Patient-Reported Outcome Measurement Information System Fatigue Scale (PROMIS-F), and the PGI-S. For screening, only the PGI-S was used to assess fatigue for enrolment. During subsequent visits (Visit 1, Visit 4, Visit 7, and Visit 10), all four PROs were recorded by participants in a paper booklet provided by the study site. Details of the schedule of study assessments are provided in the **Supplementary Materials_Table S1 and Figure S1**.

Participants also used the mAI Fatigue tool, software developed by Blueskeye® in 2023, to record AV data while performing interactive tasks on their mobile phones. The app employed machine learning algorithms to objectively analyse facial and vocal data, providing insights into expressed emotions, discomfort, stress, and fatigue. The AV tasks included a picture description activity to assess the energy of expressed emotions, a mimicking task to evaluate facial muscle movements, a read-aloud task to measure vocal muscle movements, and an eye-tracking task that involved following a moving dot and spotting a target in a cluttered image. AV recordings captured facial expressions, gaze, and voice-acoustic features, while Psychomotor Vigilance Task (PVT) reaction times were also assessed via the mobile app. All AV recordings were conducted within 30 minutes of completing the PRO assessments. Devices used for AV recordings were required to have a front-facing camera capable of capturing video in 720p resolution and a screen size of 5 inches or larger.

### Statistical Analysis

The primary endpoint analysis focused on the correlation between PRO scores and AV metrics. Correlations were assessed using statistical methods, including Pearson’s correlation (R), Spearman’s rank correlation, and Kendall’s Tau-b. To compare AV metrics between the two cohorts (CLD with fatigue vs. non-fatigued controls), appropriate statistical tests were applied based on data distribution, such as Student’s t-test, Welch’s t-test, or Mann-Whitney U test. The significance level was 5%.

To ensure consistency, PRO scores were normalised using min-max scaling, while AV metrics were standardised using Z-score normalisation. Temporal variance features and baseline corrections were applied to account for differences between participants and to address mismatches in PRO recall periods.

The per-protocol sample (PPS) was used to analyse the primary endpoint, while the full analysis set (FAS) was used for secondary analyses. As a proof-of-concept study, the target sample size of approximately 55 participants per cohort was pragmatic rather than formally powered; no a priori power calculation was performed, and all subgroup analyses are therefore exploratory. Additional subgroup analyses explored the effects of age, sex, fatigue severity, and CLD aetiology on the relationship between AV metrics and PRO scores.

## Data availability

The data that support the findings of this study are available from the corresponding author, upon reasonable request. The custom code used in this study is proprietary and consists of two parts: the biomarker extractor code and the analysis code. Analysis code can be made available upon reasonable request. Biomarker extraction can be made available as a binary executable under a suitable contractual agreement. For example, we have a standard academic use contract.

## Acknowledgements

Editorial assistance was provided by Martin Guppy PhD at Metamols Limited.

## Author contributions

All authors contributed equally to the conception, design and writing of the manuscript. All authors critically revised the manuscript, agree to be fully accountable for ensuring the integrity and accuracy of the work, and read and approved the final manuscript.

