## supplement for "Pilot Validation of an AI-based Audiovisual Fatigue Assessment Tool (mAI Fatigue) in Chronic Liver Disease: A Multicentre Study"

**Table S1.** Schedule of study assessments

| **Type of visit** | **Study site** | **At-home assessments** | | | | | | | | **Study site** |
| --- | --- | --- | --- | --- | --- | --- | --- | --- | --- | --- |
| **Visit** | **1 (screening/BL)*** | **2** | **3** | **4** | **5** | **6** | **7** | **8** | **9** | **10 (EOS)** |
| **Day** | **–2 to 1** | **3 (±1)** | **5 (±1)** | **8 (±1)** | **11 (±1)** | **13 (±1)** | **15 (±1)** | **18 (±1)** | **20 (±1)** | **22 (+2)** |
| Informed consent | X |  |  |  |  |  |  |  |  |  |
| Demographic data | X |  |  |  |  |  |  |  |  |  |
| Medical/surgical history | X |  |  |  |  |  |  |  |  |  |
| Physical examination | X |  |  |  |  |  |  |  |  | X |
| Inclusion/exclusion criteria (including PRO: PGI-S) | X^†^ |  |  |  |  |  |  |  |  |  |
| Urine pregnancy test^‡^ | X |  |  |  |  |  |  |  |  |  |
| Vital signs | X |  |  |  |  |  |  |  |  | X |
| PROs (MFI, FACIT-F, PROMIS-F, PGI-S; recorded on paper) | X**^¶^** (BL) |  |  | X |  |  | X |  |  | X |
| Dispense PRO booklet (paper) for recording PRO questionnaires at home | X (BL) |  |  |  |  |  |  |  |  |  |
| Collection of PRO booklet (recorded on paper) |  |  |  |  |  |  |  |  |  | X |
| AV recording (using a mobile tool) | X (BL) | X | X | X | X | X | X | X | X | X |
| Compliance Check |  | X^§^ | X^§^ | X^§^ | X^§^ | X^§^ | X^§^ | X^§^ | X^§^ | X |
| Prior medication | X |  |  |  |  |  |  |  |  | X |
| Concomitant medication | X |  |  | X^§^ |  |  | X^§^ |  |  | X |
| Significant event (including surgery)^\|\|^ | X (BL)^#^ |  |  | X^§^ |  |  | X^§^ |  |  | X |

*****Screening period was the period from consent till the final assessment of inclusion and exclusion criteria for eligible participants. Day 1 was considered as the day of completion of PRO and AV recording at the study site.

^†^For screening, the participants completed only PGI-S.

^‡^Urine pregnancy test was done only for child-bearing women.

**^¶^**For the baseline visit, the PGI-S score obtained at screening was used.

^§^Activities were conducted via phone calls.

^||^Significant event referred to any event, as per the investigator’s judgement, which may impact participants’ level of fatigue or participation in the study activities from screening until EOS visit.

^#^Significant event at baseline includes any event identified after consenting and post-enrolment.

AV, audiovisual; BL, baseline; EOS, end of study; FACIT-F, functional assessment of chronic illness therapy fatigue scale; MFI, multidimensional fatigue inventory; PGI-S, patient global impression of severity; PRO, patient-reported outcome; PROMIS-F, patient-reported outcome measure information system for fatigue.

**Table S2.** Participant disposition

| **Characteristics** | **Cohort A (N=55) n (%)** | **Cohort B (N=56) n (%)** |
| --- | --- | --- |
| Number of participants who completed the study | 54 (98.2) | 50 (89.3) |
| Number of participants who prematurely terminated the study | 1 (1.8) | 6 (10.7) |
| Primary reason for premature study termination |  |  |
| Dropouts | 1 (1.8) | 6 (10.7) |
| Significant event | 0 | 0 |
| Urgent surgery of fatigue medication needs | 0 | 0 |
| Lost to follow-up | 0 | 3 (5.4) |
| Withdrawal of consent | 1 (1.8) | 2 (3.6) |
| Protocol violation | 0 | 1 (1.8) |
| Sponsor request | 0 | 0 |
| Physician decision | 0 | 0 |
| Other | 0 | 0 |

**Table S3.** Chronic liver disease diagnosis and aetiology (all subjects enrolled)

| **Characteristics** | **Cohort A (N=55) n (%)** | **Cohort B (N=56) n (%)** |
| --- | --- | --- |
| CLD diagnosis |  |  |
| Yes | 0 | 56 (100) |
| No | 55 (100) | 0 |
| CLD aetiology |  |  |
| Viral hepatitis B | 0 | 0 |
| Viral hepatitis C | 0 | 1 (1.8) |
| Viral hepatitis D | 0 | 0 |
| Viral hepatitis E | 0 | 4 (7.1) |
| Metabolic dysfunction-associated steatotic liver disease (MASLD) | 0 | 33 (58.9) |
| Non-alcoholic steatohepatitis (NASH) | 0 | 1 (1.8) |
| Autoimmune hepatitis | 0 | 0 |
| Primary biliary cholangitis | 0 | 1 (1.8) |
| Primary sclerosing cholangitis | 0 | 0 |
| Genetic disorder | 0 | 0 |
| Metabolic disorder | 0 | 0 |
| Alcoholic liver disease | 0 | 16 (28.6) |

CLD, chronic liver disease.

**Table S4.** Audiovisual markers

| **Marker category** | **Markers** |
| --- | --- |
| Blink dynamics | Eye Lid Metric 1–33 |
| Gaze behaviour | Gaze Metric 1–18 |
| Voice acoustics | Audio Metric 1–55 |
| Facial action units (AUs) | AU Metric 1–66 and 73-86 |
| Mouth movements | Mouth Metric 1–6 |

AU, action unit.

**Table S5.** Manual audiovisual quality annotation results for audiovisual tasks

| AV Task | Visual quality | | | Audio quality | | |
| --- | --- | --- | --- | --- | --- | --- |
|  | **Total valid** | **Good** | **Bad** | **Total valid** | **Good** | **Bad** |
| **Gaze reaction** | 396/400 | 181 (45.7%) | 215 (54.3%) | Not Applicable | Not Applicable | Not Applicable |
| **Read aloud** | 390/400 | 198 (50.8%) | 192 (49.2%) | 390/400 | 142 (36.4%) | 248 (63.6%) |
| **Picture description** | 384/400 | 118 (30.7%) | 266 (69.3%) | 384/400 | 75 (19.5%) | 309 (80.5%) |

AV, audiovisual.

**Table S6.** Audiovisual markers with high consistency ranking

| Filter Type / Feature Type | Blink | Gaze | Voice | AUs | Mouth |
| --- | --- | --- | --- | --- | --- |
| None | Eye Lid Metric 27 | Gaze Metric 12  Gaze Metric 2  Gaze Metric 1  Gaze Metric 9  Gaze Metric 16 | Audio Metric 35  Audio Metric 13  Audio Metric 20 | AU Metric 27  AU Metric 79  AU Metric 15  AU Metric 25  AU Metric 44  AU Metric 2 | - |
| Visual Quality | Eye Lid Metric 1  Eye Lid Metric 19  Eye Lid Metric 2  Eye Lid Metric 4  Eye Lid Metric 7  Eye Lid Metric 14  Eye Lid Metric 26  Eye Lid Metric 8  Eye Lid Metric 22  Eye Lid Metric 23  Eye Lid Metric 29 | Gaze Metric 12  Gaze Metric 16  Gaze Metric 11  Gaze Metric 10  Gaze Metric 18 | Audio Metric 2  Audio Metric 3  Audio Metric 18  Audio Metric 4  Audio Metric 31 | AU Metric 25  AU Metric 31  AU Metric 34  AU Metric 79  AU Metric 15  AU Metric 27  AU Metric 36 | - |
| Audio Quality | Eye Lid Metric 6  Eye Lid Metric 17  Eye Lid Metric 12  Eye Lid Metric 2 | Gaze Metric 12  Gaze Metric 16  Gaze Metric 8  Gaze Metric 6  Gaze Metric 7 | Audio Metric 4  Audio Metric 33  Audio Metric 20  Audio Metric 13  Audio Metric 38  Audio Metric 47 | AU Metric 31  AU Metric 58  AU Metric 9  AU Metric 16  AU Metric 18  AU Metric 36  AU Metric 66  AU Metric 7  AU Metric 49 | Mouth Metric 5  Mouth Metric 4 |

**Table S7.** Correlation directions between audiovisual fatigue markers and patient-reported outcome measures by severity of fatigue group (combined cohorts, N=104)

| **Task** | **PRO score** | **AV marker** | **Direction** | **None** | **Moderate** | **Severe** |
| --- | --- | --- | --- | --- | --- | --- |
| Picture description | MFI Physical Fatigue | AU Metric 83 | Negative correlation |  | rho: –0.4699  P: 0.0003  N: 41 | rho: –0.5545  P: 0.0209  N: 9 |
| Picture description | MFI Mental Fatigue | Audio Metric 1 |  | rho: 0.1505  P: 0.0367  N: 50 | rho: –0.2233  P: 0.0047  N: 41 |  |
| Eye tracking | MFI Physical Fatigue | AU Metric 75 |  | rho: 0.2136  P: 0.0956  N: 50 |  | rho: –0.8964  P: 0.0063  N: 9 |

AU, action unit; AV, audiovisual; MFI, multidimensional fatigue inventory; PRO, patient-reported outcome.

**Table S8.** Correlation of fatigue model with patient-reported outcome measures (Good Visual Quality subset, N=55)

|  | **RMSE** | **Pearson correlation coefficient (R)** |
| --- | --- | --- |
| **Correlation with MFI Physical Fatigue** | 0.4360 | 0.3850 |
| **Correlation with MFI Mental Fatigue** | 0.3845 | 0.5344 |

MFI, multidimensional fatigue inventory; RMSE, root mean squared error.

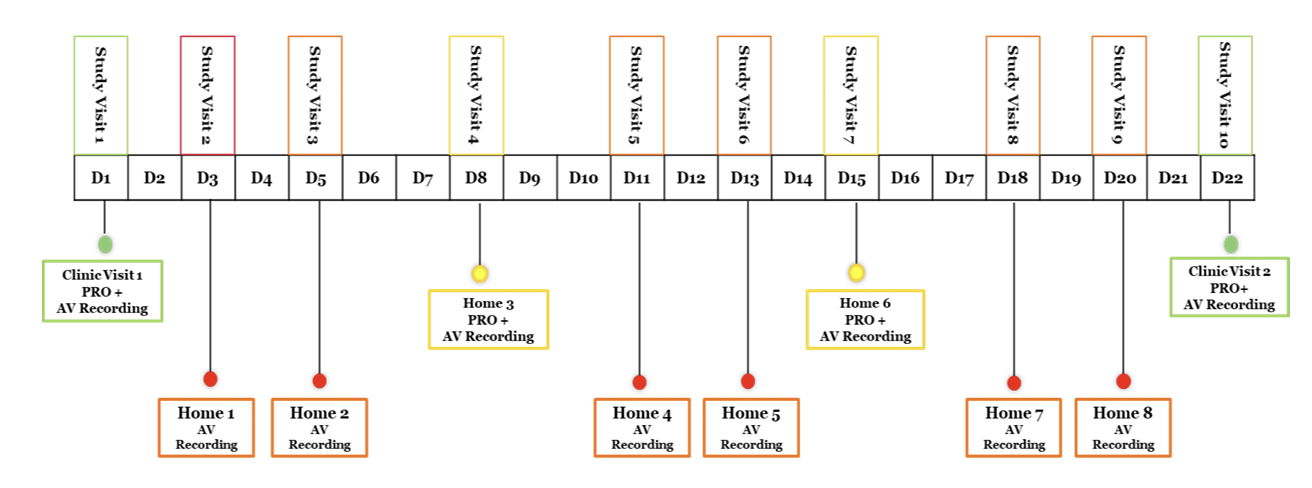
**Figure S1.** Schedule of study assessments

AV, audiovisual; D, day; PRO, patient-reported outcome.

**Figure S2.** Participant disposition

**N=116**

Subjects consented

**N=111**

Enrolled

**N=5**

Screen failures

**N=55**

Cohort A

**N=56**

Cohort B

**N=54**

Completed

**N=1**

Terminated

prematurely

**N=50**

Completed

**N=6**

Terminated

prematurely

**Figure S3.** Trend consistency analysis plots for four distinct candidate fatigue markers

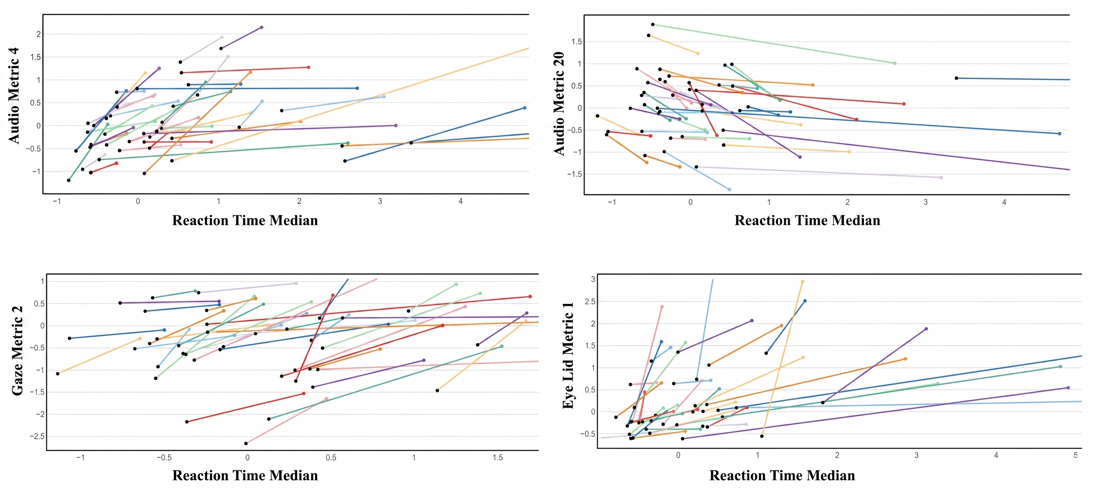

Note: Each coloured line represents an individual participant, tracking the change in their specific marker value as they progress from their lowest to highest fatigue state (proxied by Reaction Time Median). A strong relationship is indicated when the majority of participant lines follow a consistent directional slope.

**Figure S4**. Correlation and distribution plots for top participant-level features (Good Visual Quality subset, N=55) vs. MFI Physical Fatigue

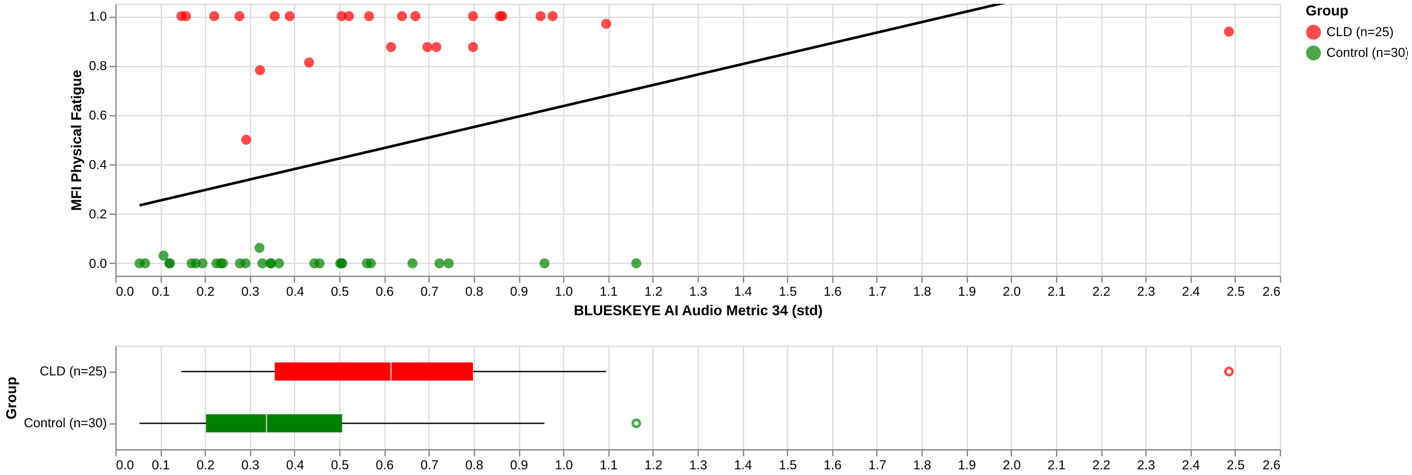

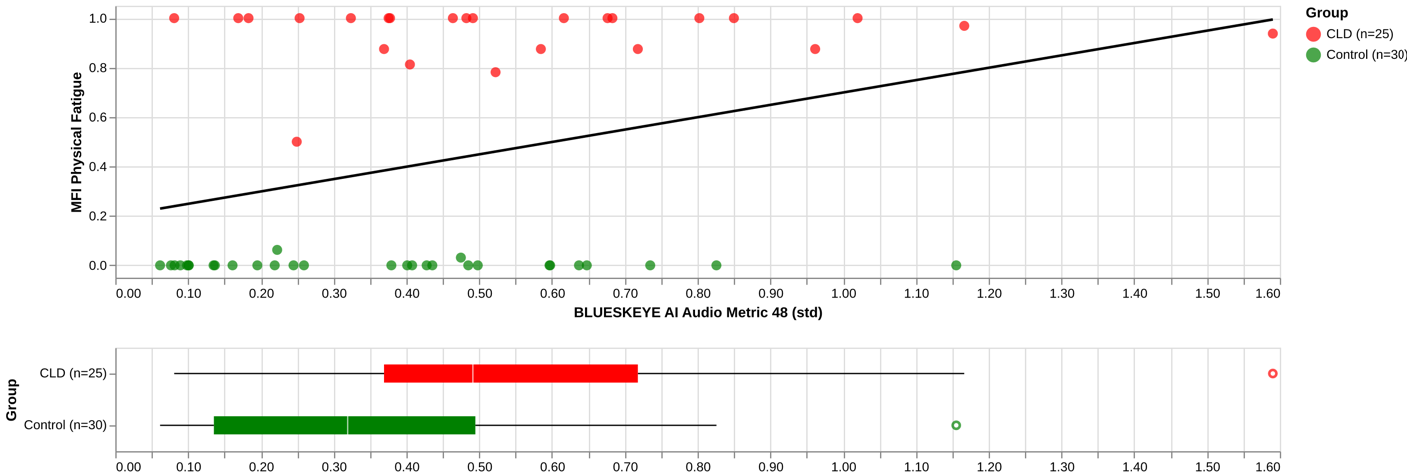

Note: The features displayed on the X-axis (labeled ‘SD’) represent the temporal standard deviation of an AV metric across all available PRO visits of a participant. This captures the variability of AV metrics as a predictor of fatigue.

**Figure S5**. Correlation and distribution plots for top participant-level features (Good Visual Quality subset, N=55) vs. MFI Mental Fatigue

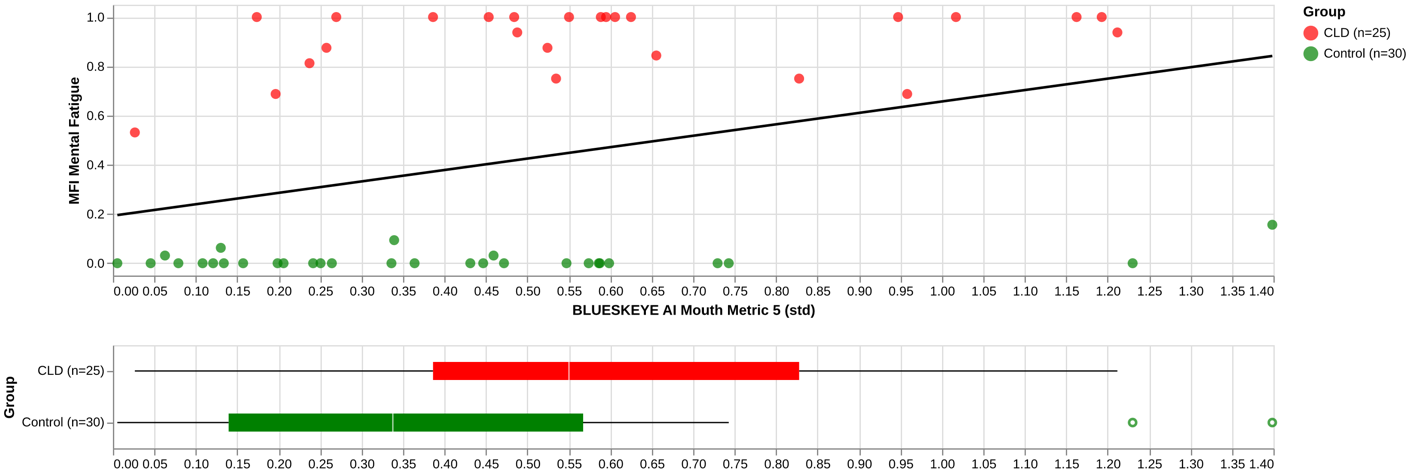

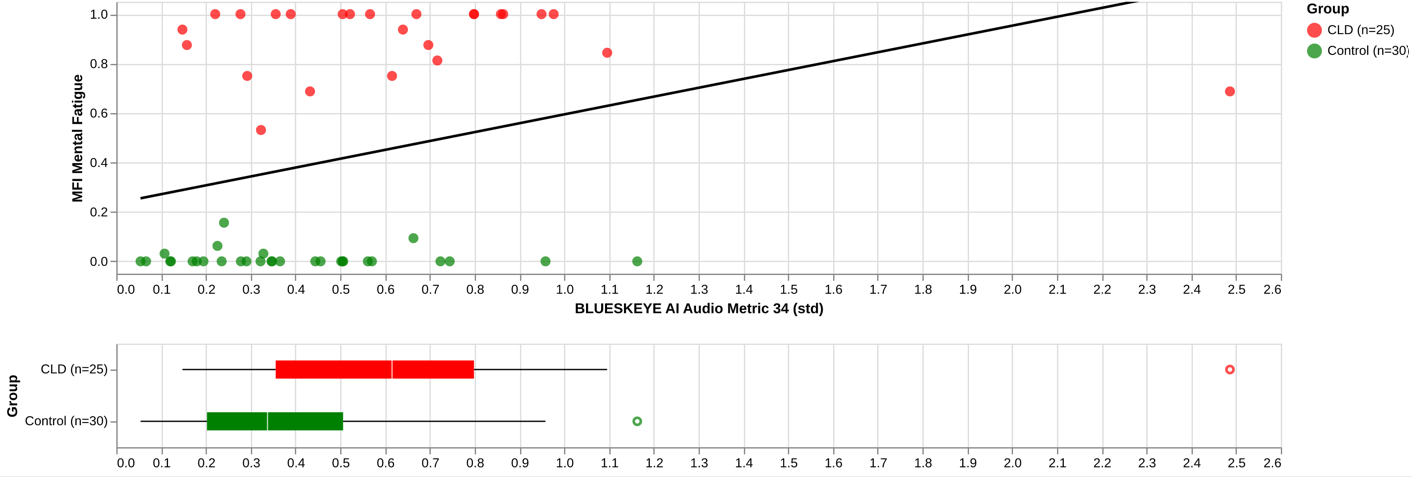

Note: The features displayed on the X-axis (labeled ‘SD’) represent the temporal standard deviation of an AV metric across all available PRO visits of a participant. This captures the variability of AV metrics as a predictor of fatigue.

**Figure S6.** Correlation of individual audiovisual features vs. fatigue model outputs with MFI Physical Fatigue (Good Audio Quality subset, N=41)

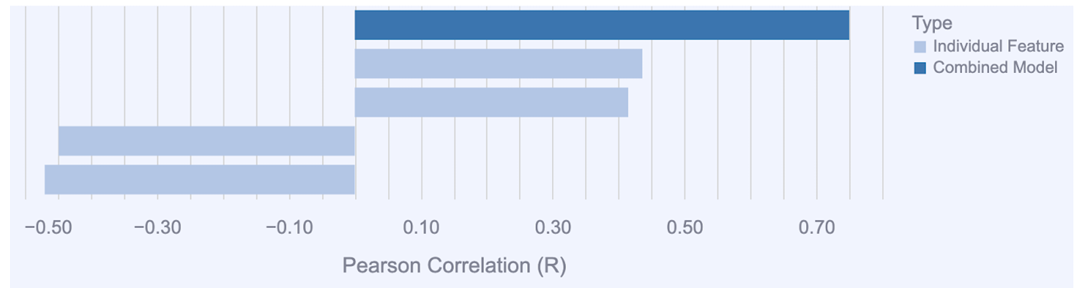

Audio Metric 22

Eye Lid Metric 28

Mouth Metric 4
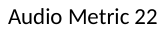

Eye Lid Metric 18

MFI, multidimensional fatigue inventory.

**Figure S7.** Correlation of individual audiovisual features vs. fatigue model outputs with MFI Physical Fatigue (Good Visual Quality subset, N=55)

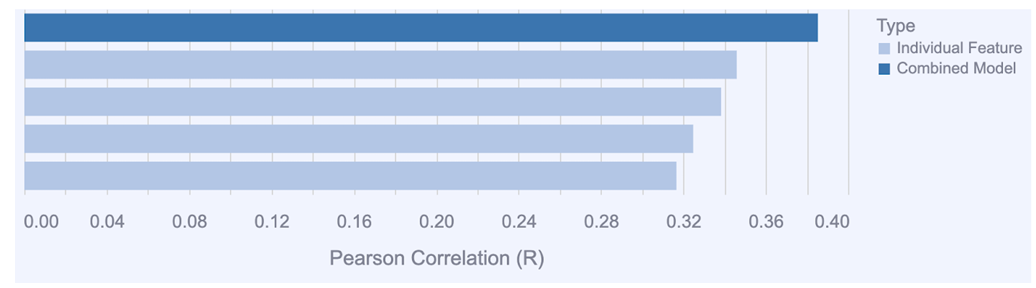

Audio Metric 34

Audio Metric 48

Audio Metric 43

Audio Metric 40

MFI, multidimensional fatigue inventory.

**Figure S8.** Predicted vs. actual fatigue scores for MFI Physical Fatigue (Good Visual Quality subset, N=55)

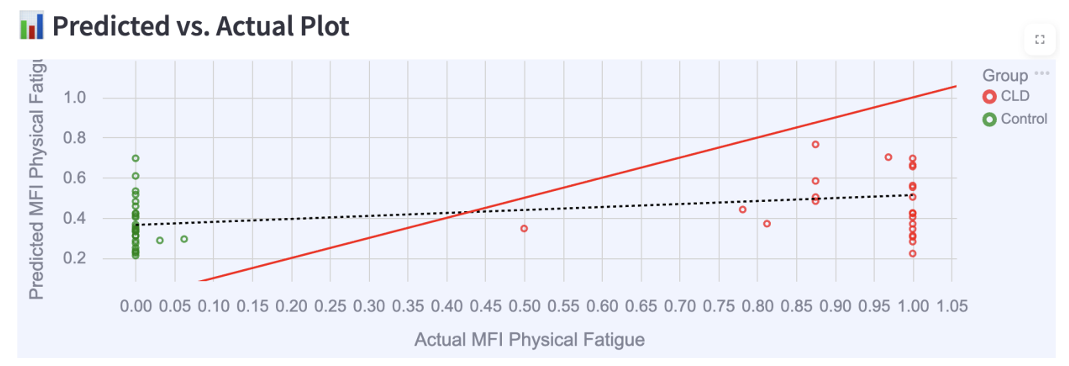

Predicted MFI Physical Fatigue

Note: The solid red line indicates the ideal reference (y=x) representing perfect prediction. The dotted black line indicates the linear best-fit regression of the model’s predictions. The vertical distance between a point and the red line represents the prediction error for that participant.

CLD, chronic liver disease; MFI, multidimensional fatigue inventory.

**Figure S9.** Correlation of individual audiovisual features vs. fatigue model outputs with MFI Mental Fatigue (Good Visual Quality subset, N=55)

Audio Metric 34

Eye Lid Metric 21
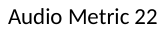

Eye Lid Metric 20

Mouth Metric 4

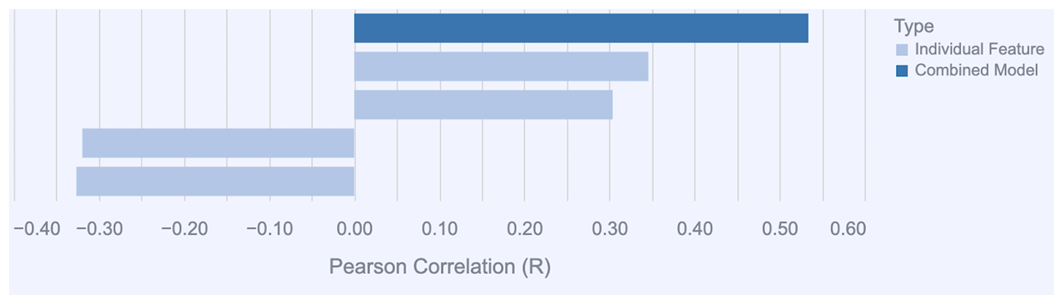
MFI, multidimensional fatigue inventory.

**Figure S10.** Predicted vs. actual fatigue scores for MFI Mental Fatigue (Good Visual Quality subset, N=55)

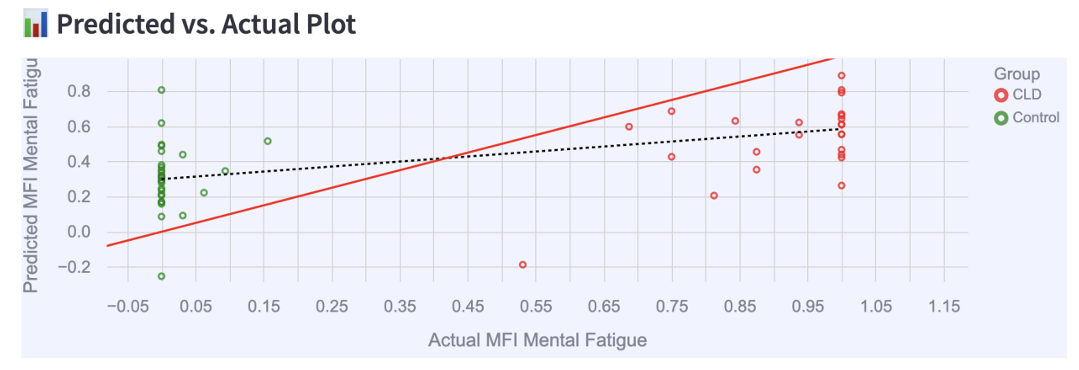

Predicted MFI Mental Fatigue

Note: The solid red line indicates the ideal reference (y=x) representing perfect prediction. The dotted black line indicates the linear best-fit regression of the model’s predictions. The vertical distance between a point and the red line represents the prediction error for that participant.

CLD, chronic liver disease; MFI, multidimensional fatigue inventory.
